# Identification of the molecular components of enhancer-mediated gene expression variation in multiple tissues regulating blood pressure

**DOI:** 10.1101/2023.12.07.23299084

**Authors:** Or Yaacov, Prabhu Mathiyalagan, Hanna H. Berk-Rauch, Santhi K. Ganesh, Luke Zhu, Thomas J. Hoffmann, Carlos Iribarren, Neil Risch, Dongwon Lee, Aravinda Chakravarti

## Abstract

Inter-individual variation in blood pressure (BP) arises in part from sequence variants within numerous enhancers modulating expression of an unknown number of causal genes. We propose that these genes are active in tissues relevant to BP physiology and can be identified from tissue epigenomic data and genotypes of BP-phenotyped individuals. We used the ENCODE project’s H3K27ac and ATAC-seq data from the heart, adrenal gland, kidney, and artery to comprehensively identify all cis regulatory elements (CREs) in these tissues to estimate the impact of all common human single nucleotide variants (SNVs) in CREs on gene expression, using machine learning methods. To identify specific genes, we integrated these results through a gene-wise association test against BP. We conducted analyses in two separate large-scale cohorts: 77,822 individuals from the Genetic Epidemiology Research on Adult Health and Aging (GERA) of Kaiser Permanente North California and 315,270 individuals from the UK Biobank (UKB).

We identified 309, 259, 331 and 367 genes (FDR<0.05) for diastolic BP (DBP), and 191, 184, 204, and 204 genes for systolic BP (SBP), in the artery, kidney, heart, and adrenal gland, respectively, in GERA; 50-70% of these genes were replicated in the UKB and is significantly higher than the 12-15% expected by chance (P <10^-4^). These results enabled the prediction of tissue expression of these 988-2,875 putative BP-genes in individual participants of both cohorts to construct an expression polygenic score (exPGS). This score explained ∼27% of the reported SNV heritability (h^2^, 21%), substantially higher than that expected from prior studies. Additionally, we utilized these methods to provide dual-modality supporting evidence, CRE and expression-based, for the causality of genes previously detected by GWAS.

## INTRODUCTION

Although the physiology of blood pressure (BP) is well understood, its genetic control is largely elusive. BP is a classical quantitative trait and the subject of long debate regarding its inheritance: is its inter-individual distribution continuous (Pickering) or discrete (Platt), that is, is its genetic architecture polygenic or monogenic?^1^ It is now well established that systolic (SBP) and diastolic (DBP) blood pressure have continuous distributions across humans, have recognizable age- and sex-specific trends, with little distributional difference between world-wide populations.^2^ BP is a textbook example of a polygenic trait with 30%-50% heritability in adults but also subject to variation from many environmental and lifestyle factors.^3^

The largest BP genomic investigation, using genome-wide association studies (GWAS) in more than 1 million participants, overwhelmingly demonstrated that the majority of its heritability is explained by ∼900 genetic associations of single nucleotide variants (SNVs) of small effects distributed across the non-coding genome.^4^ Further, this study estimated that the SNV-wide heritability (h^2^) is ∼0.21 for both SBP and DBP. Thus, the previously known genes with major monogenic effects leading to hypertension and hypotension syndromes,^5^ and affecting renal and adrenal physiology through salt-water homeostasis, are mechanistically important but more of an exception than the rule in inter-individual population BP variation. The combined conclusion from these studies is that variation in BP traits largely arises from numerous common regulatory variants (see below) each of small effect in numerous genes acting through multiple tissues, but that rare pathogenic protein function-altering variants in specific renal and adrenal genes can have large impact on BP in rare individuals. Thus, extreme BP values, relevant to hypertension and hypotension, arise as outliers of either polygenic or monogenic variation. Which of these classes are more frequent in BP extremes, and its target organ damages when untreated, is, however, unresolved and an important question to answer in human physiology.

In this study we tackle this question by identifying the *specific* genetic, genomic and tissue components that underlie polygenic contributions to BP variation. Blood pressure GWAS, similar to other complex traits, have implicated thousands of SNVs contributing to the polygenic BP spectrum but their molecular routes of action are unknown because the majority of these variants are noncoding and include both causal and passenger (in linkage disequilibrium (LD) with causal) variants.^4^ Other studies have estimated that >75% of significantly associated noncoding GWAS variants are located within cis-regulatory elements (CREs) or are in high LD with CREs,^6^ and, therefore, believed to carry their effects through regulation of gene expression. Transcription factors (TF) bind these CREs and modulate the transcription of BP causal genes in specific tissues, as CREs and their cognate TFs vary by cell type and tissues. Thus, a tissue-agnostic approach to understanding polygenic BP is unsuitable because the regulatory genomic components are not universal across tissues.^7,8^ A genetic dissection of polygenic BP entails identifying the CREs, their cognate TFs and the CRE SNVs that change CRE activity on a tissue-by-tissue basis. These results lead to definitive gene identification, enabling screening these genes for rare variants with monogenic effects.

Our previous work^8^ demonstrated that regulatory variants specific to the artery have the largest (11.8%) contribution to BP heritability, followed by those from the heart (7.7%) and adrenal (5.3%); surprisingly, the kidney-specific CRE variants had the lowest contribution (2.5%), consistent with prior GWAS data.^7,9,10^

In this study, we use machine learning methods^11–13^ we previously developed to construct comprehensive genome-wide chromatin accessibility maps on surgical and/or cadaverous tissues of independent individuals, and biobank scale genotypes of BP-phenotyped individuals^9,14^ to detect and validate specific genes that regulate BP in specific tissues (**Figure 1**).

**Figure 1:**
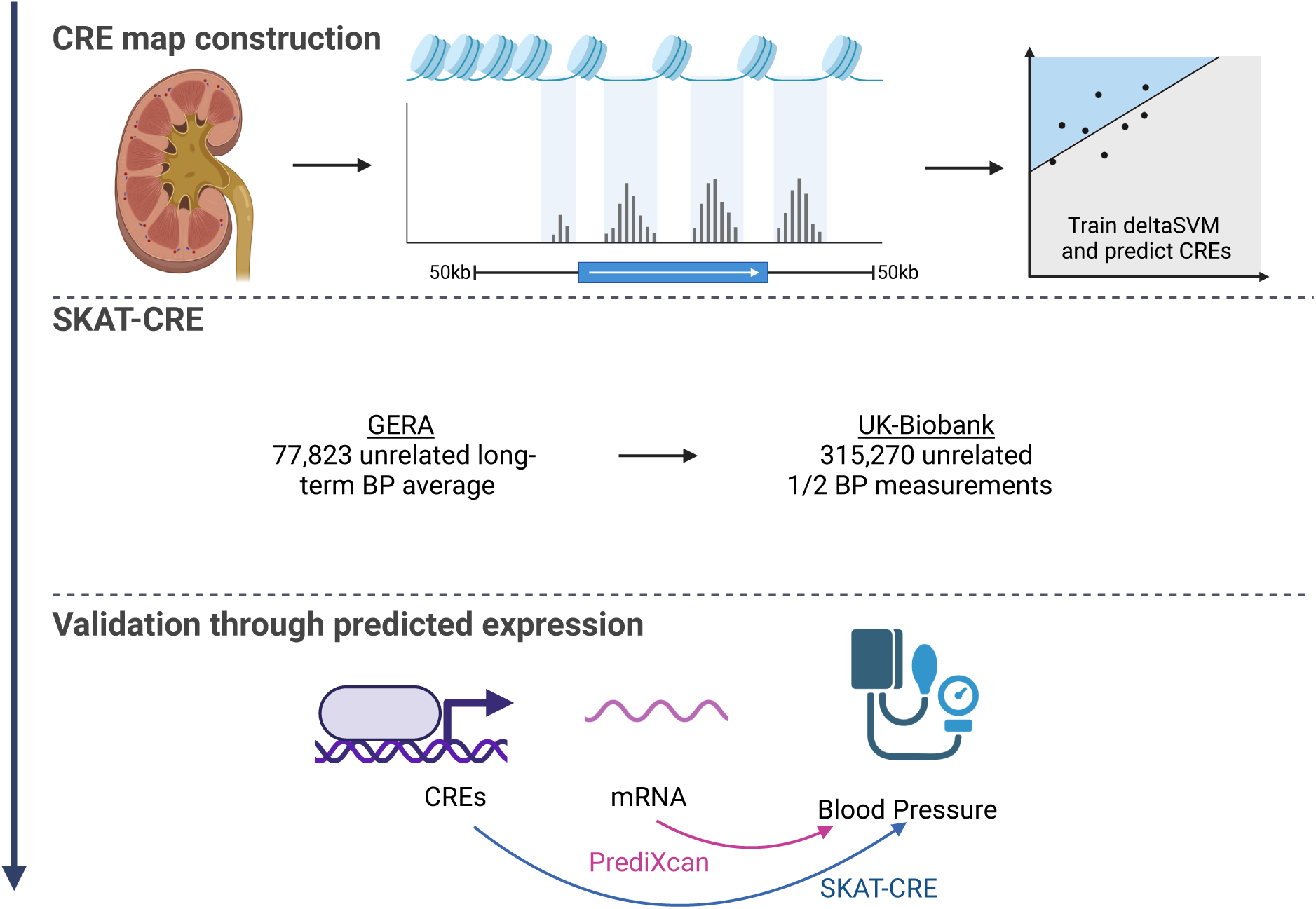
Three-step study design for complex trait regulatory genomic analyses using cis regulatory element (CRE) map construction from individual tissues (top panel, kidney given as an example), modified SKAT association testing using the identified CREs (SKAT-CRE analysis) on Biobank scale (GERA, UKB) datasets (middle panel), and validating the identified genes through their gene expression variation effects on a complex trait (e.g., BP). This design is applied to each of the tissues separately.

## RESULTS

### Constructing BP tissue-restricted epigenomic maps

For this study, we included four BP-relevant tissues, namely, the heart (left ventricle), adrenal gland, kidney (cortex), and (tibial) artery, that we have recently constructed. The maps for heart, adrenal gland, and artery were generated from publicly available DNase-seq and ATAC-seq data from the ENCODE project.^15^ For the kidney, our laboratory generated ATAC-seq data from 4 samples, owing to the paucity of publicly available adult human kidney open chromatin maps. Human samples of the same tissue vary in the peaks they capture as a result of technical and biological factors, as the tissues were procured from different donors.^8^ We, therefore, created one comprehensive map for each tissue, using the best-performing CREs as determined by our quality assessment using gkmQC.^16^ The final numbers of CREs that passed our quality control parameters, as described in the Methods section, and based on our previous work,^8^ are 141,510, 133,231, 132,970, 104,812 in the adrenal, kidney, heart, and tibial artery, respectively. These CREs in these tissues cover 12.2%, 11.4%, 13.6% and 11.1% of the genome, respectively. Collectively, the CREs from all of the four tissues cover 24.3% of the genome. These maps are available in the Supplementary material.

### Identifying CRE regulatory variants

Tissue-restricted epigenomic maps were further annotated by sequence-based regulatory variant prediction. We calculated a score for each common variant (minor allele frequency (MAF) ≥1%) from the 1000 Genomes Project^17^ residing within CREs using the deltaSVM^18^ method (see Methods). A total of 1,253,435, 1,183,563, 1,414,660 and 1,139,136 variants mapped within the CREs of the adrenal, kidney, heart and artery, respectively. These variants are the subsets of all 10,041,373 common variants in the genome (see Methods) which fall within CREs, and their deltaSVM scores represent their predicted regulatory activity. These numbers are consistent with previous works.^7,8^ The result is a tissue-restricted map of common variants in CREs and their associated impact on chromatin accessibility in that tissue as measured by deltaSVM scores.

### Identifying tissue-restricted BP genes using SKAT-CRE association tests

For studies of rare variants of a gene impacting a trait, the sequence kernel association test (SKAT)^19^ is an efficient method to combine the effects of multiple rare variants by weighting variants inversely proportional to their allele frequency. We adapted this test for assessing the contributions of all common variants in all CREs for a target gene, which we refer to as SKAT-CRE, to detect CRE associations with SBP and DBP. Conventionally, in SKAT, common variants with MAF>1% are usually weighted close to zero. Instead, in SKAT-CRE, we weighted variants by their functional impact as predicted by their deltaSVM scores in a tissue-restricted way. These analyses were conducted for each tissue separately.

We conducted a gene-wise SKAT-CRE analysis for each protein-coding gene (GENCODE v19).^20^ We only included genes expressed in a tissue of interest with a median expression of at least one transcript per million (TPM) across GTEx v8^21^ samples from that tissue, a conventional GTEx expression threshold. This gene selection scheme resulted in similar-sized sets of 12,478, 12,412, 12,353, and 11,058 genes in the kidney, artery, adrenal gland, and heart, respectively. CREs were defined within 50 kb (‘cis’) of the gene start or end, including the gene body using GENCODE v19 gene annotations.^20^ The boundary was determined based on the observation that more than 85% of lead eQTLs are located within 50 kb of the gene.^22^ All common SNVs within these CREs, deltaSVM score weighted as indicated, were considered putative regulatory variants. Only variants that appear in the relevant tissue CRE map were included in each tissue-restricted analysis. The number of CREs per gene, defined as the gene body ±50 kb, as well as regulatory variants within the CREs was also similar across the four tissues (**Figure 2 A, C**). In each tissue, 11%-24% of the CREs were unique to the tissue, 34%-41% were common to 2-3 tissues while 39%-49% were ubiquitous (**Figure 2B**). Expectedly, the number of CREs per gene was significantly correlated (r = 0.54-0.61) with its gene length (**Supplementary Figure S1**).

**Figure 2:**
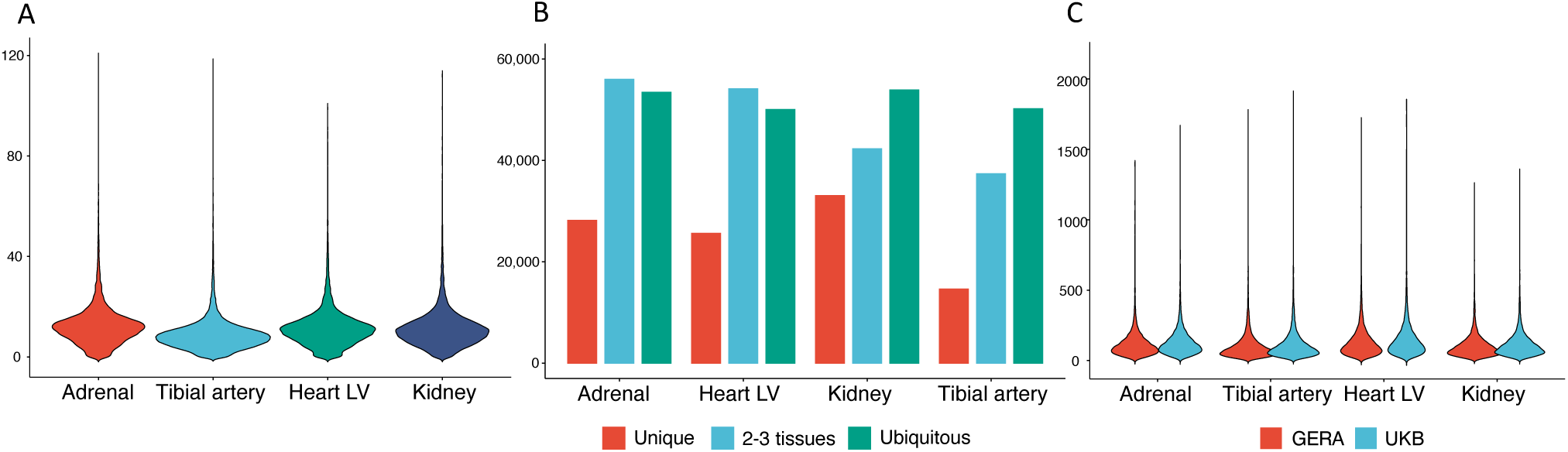
Statistical properties of CREs and their regulatory sequence variants. **A.** Distribution of the numbers of CREs per gene in each of the four tissues studied. **B.** Numbers of CREs unique to one tissue (red), common to 2-3 tissues (blue) or ubiquitous across all 4 tissues (teal). **C.** Distribution of the numbers of regulatory variants (within CREs) for each gene analyzed in the GERA (red) and the UKB (blue) cohorts.

BP residuals calculated for each participant and genotypes for each putative CRE variant were obtained from directly genotyped or imputed variants. In the discovery dataset (GERA), we identified 191, 184, 204, and 204 genes to have a statistically significant association with SBP in the artery, kidney, heart, and adrenal gland (Figure 3A), after adjustment for multiple comparisons using the Benjamini-Hochberg method at a false discovery rate (FDR) of 0.05. Quantile-quantile plots of nominal P values are provided in **Supplementary Figure S2**. Similarly, we detected 309, 259, 331 and 367 genes for DBP. Of these genes, 50-70% were replicated with the same statistical significance in the larger and independent UKB dataset of 315,270 unrelated participants (**Figure 3B**). The methods used to analyze the UKB data were identical to those used for GERA and yielded a larger number of significant genes (**Figure 3**). This is expected given the larger cohort size of UKB. To compare these numbers to those expected by chance, we performed permutation analysis by randomly sampling the same number of genes 100,000 times to estimate the expected replication rate: this was 12-15%, significantly lower than the observed rate of 50%-70% (**Figure 3**). The difference between the expected and observed rates of replicated genes is highly significant across all tissues and phenotypes (P<10^-4^). In the GERA cohort, 21%-29% of the significant genes detected in each tissue were common to SBP and DBP. In the UKB, 53%-55% of the detected genes in each tissue were common to SBP and DBP.

**Figure 3:**
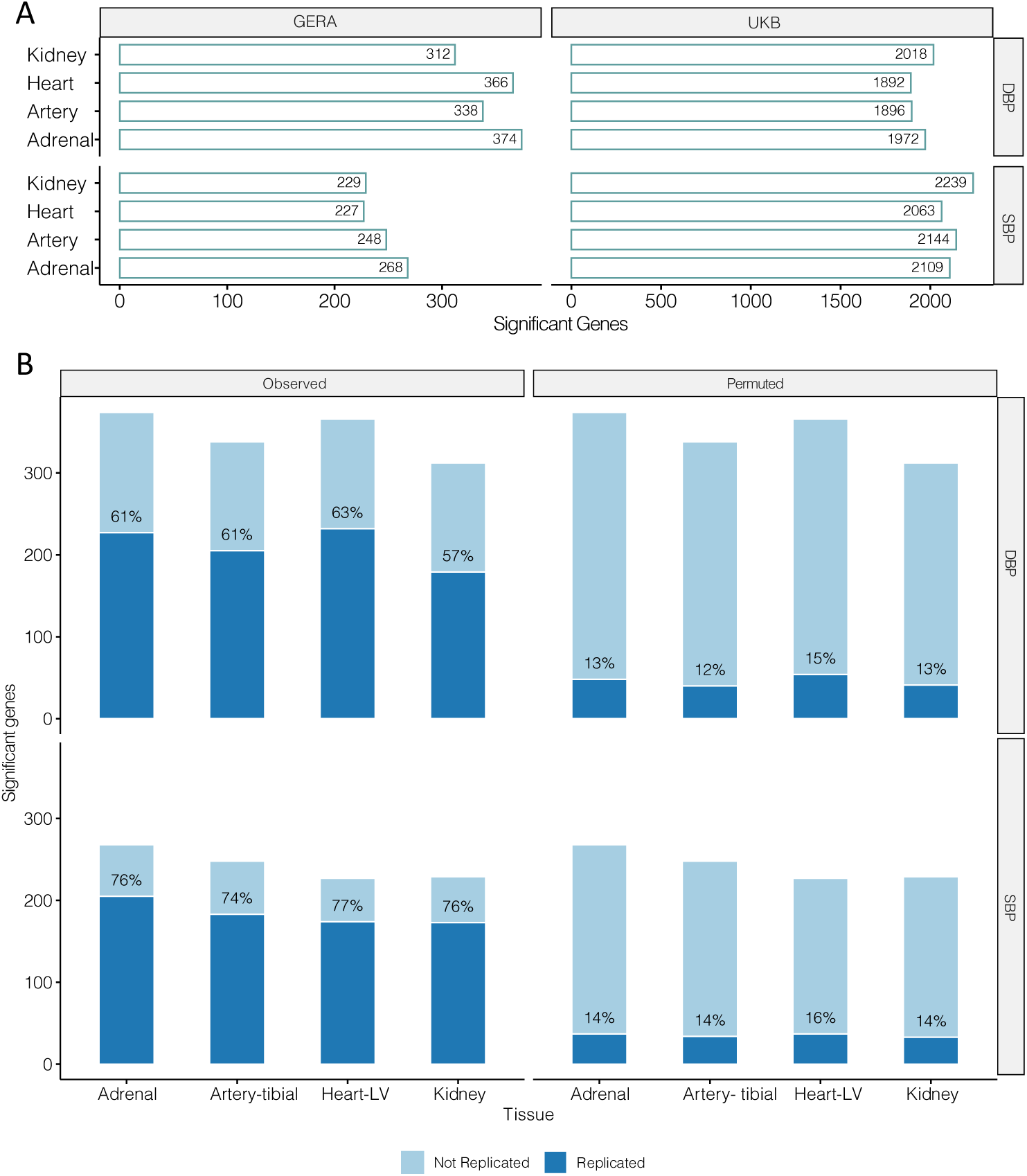
SKAT-CRE association study. **A.** Numbers of statistically significant genes (Type I error of 0.05, Benjamini-Hochberg false discovery rate (FDR) adjusted) detected in each tissue in the discovery (left) and replication (right) cohorts. **B.** The numbers and proportions of statistically significant genes replicated in the SKAT-CRE analysis (observed) versus their null expectation (by permutation analysis). DBP/SBP are diastolic/systolic blood pressure.

For the 3,220 and 3,073 unique genes detected in the SKAT-CRE analysis for SBP and DBP in either cohort, we searched the literature for known associations with SBP or DBP. We used summary statistics from the largest BP GWAS to date by Evangelou et al.^4^ which includes over 1 million participants. We searched these GWAS results for genes in proximity (50 kb) to a genome wide significant SNP (P < 5×10^-8^): 2,962 SBP and 3,211 DBP genes were found in these summary statistics. Of these genes, 1,564 SBP and 1,645 DBP genes overlapped the SKAT-CRE genes detected in our study. These numbers represent 48.6% and 53.5% of the totals we identified in our analysis. In addition, we searched the GWAS catalog (v1.0.2)^23^ for associations that were not reported by Evangelou et al.^4^ but previously reported as significant in other smaller GWAS. This search identified an additional 22 SBP and 13 DBP genes in the SKAT-CRE analysis as previously reported associations. The remaining 1,634 SBP and 1,415 DBP genes detected in our analysis were not previously reported to be associated with BP.

### Enhancer variation affects BP through gene expression variation

The significant genes detected in the SKAT-CRE analysis above arise from association of CRE genotypes with SBP and DBP. Consequently, we wanted to directly test whether these enhancers affect BP through gene expression variation in the corresponding tissues. Ideally, this test requires the gene expression profile of each of the study participants in each tissue. Since measured tissue expression data is unavailable for living participants, we alternatively used the predicted expression using LASSO regression models trained on GTEx^21^ data per gene, as implemented by PrediXcan.^24^

In order to have a comparable set of CREs across different analyses, we sought to predict gene expression using only variants in the CREs used for SKAT-CRE analysis. We constructed our own LASSO-regression gene expression models from CRE-restricted genotypes, using individual level genotypes from GTEx V8,^21^ which we refer to as CRE-restricted models: their properties are presented in **Supplementary Figure S3**. We followed previously established criteria for model significance (see Methods). The important distinction between our models and the general PrediXcan models, which we refer to as all-variant models, is that only variants in CREs active in the corresponding tissue are included in our models. Even though CRE-restricted models used only a subset of variants for model training, we found that their predictive power is similar, and at times even greater, in detecting statistically significant models for most genes. Moreover, there is low (15∼20%) overlap of included variants between the two models (**Supplementary Figure S4**). This is partly because variants outside CREs in high LD with functional variants within CREs tend to be selected first in the original models. Most of the significant models were significant in both model types but the CRE-restricted type had a slightly higher number of significant models than the all-variant type (**Supplementary Figure S5**). The number of variants available for LASSO regression (variants in window) was 5 to 7 fold larger in the all-variant models, and the final set of variants included in the model (variants in model) was slightly higher in the all-variant models as well, but with similar significance levels overall (**Supplementary Figure S6 and S7**). We compared the expression levels predicted by the different types of models using the GERA genotype data to discover that the Pearson correlation coefficient is ≥0.75 for almost all genes (**Supplementary Figure S8**). In sum, our CRE-restricted models achieved similar power and significance but from using a smaller set of putative regulatory variants in each tissue.

Next, we used the gene expression predicted by the CRE-restricted models to estimate the correlation of each gene with SBP and DBP, as implemented in PrediXcan. We performed the analysis in both the GERA and the UKB cohorts. For each gene, we compared the P values (for SBP, DBP) in the SKAT-CRE analysis to the P values in the PrediXcan analysis (SBP, DBP). In each of the four tissues, these were significantly correlated (**Figure 4**) further supporting our findings.

**Figure 4:**
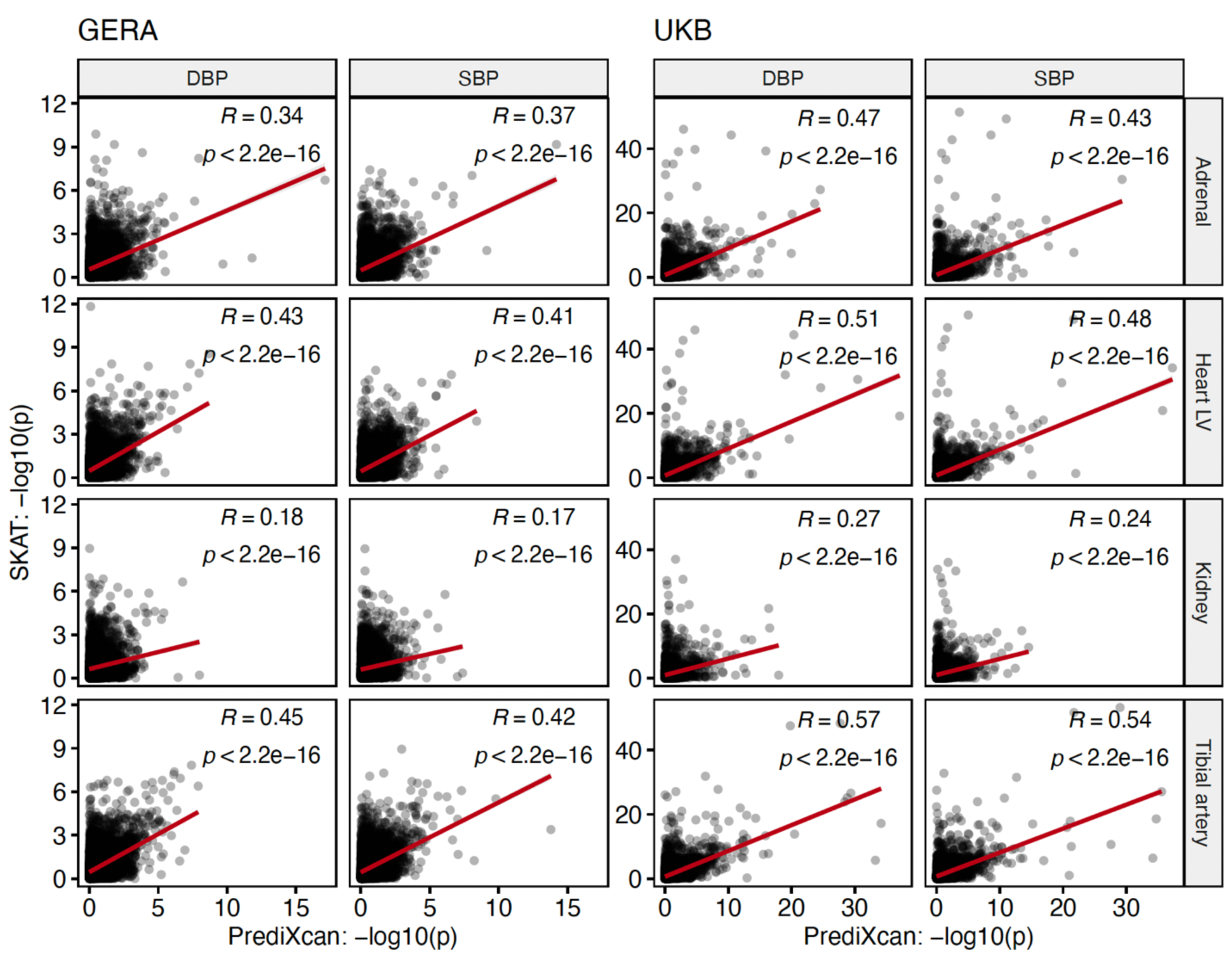
Correlations between the -log_10_ significance values of SKAT-CRE and PrediXcan association test statistics in UKB (left) and GERA (right) cohorts. DBP/SBP are diastolic/systolic blood pressure.

### Tissue-restricted gene expression predicts BP traits

Given the statistical significance of the association between predicted tissue-restricted gene expression and BP (PrediXcan), and the CRE genotypes with BP (SKAT-CRE), we sought to show that BP phenotypic variation can be explained by the predicted expression of tissue-restricted genes we identified. In order to quantify the aggregate effect of tissue CRE dependent gene expression on BP, we constructed an expression-based polygenic score (exPGS), separately for each tissue, containing only the genes detected in that tissue. Additionally, we also estimated for each study participant a combined all-tissue exPGS.

We first constructed a summation-based exPGS for each set of genes, as follows:

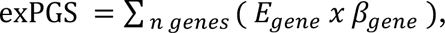

as a sum of the normalized predicted expression (*E_gene_*) of each gene weighted by its effect size (*β_gene_*) from the PrediXcan association test described above. The combined all-tissue exPGS was defined as the arithmetic mean of all 4 tissue exPGS scores per participant. To demonstrate the performance and specificity of exPGS, we tested the correlation between exPGS values as binned percentiles to BP, as well as to an unrelated control phenotype BMI. As shown in **Figure 5**, exPGS percentiles were significantly correlated with SBP (r=0.948, P<2.2×10^-16^), but not with BMI (r=-0.16, P=0.2). The results were similar for DBP (r=0.949, P<2.2×10^-16^) and were consistent across both the GERA and UKB cohorts. In order to limit the possibility of overfitting, we further tested the exPGS derived from genes detected in one cohort on the genotypes and phenotypes of the other cohort. We next calculated the Pearson correlation coefficient between all possible exPGS scores and the phenotype (**Table 1**). These analyses show that the exPGS based on GERA genes had Pearson correlation coefficients of 0.06∼0.09 for the four tissues and 0.1 for all tissues combined when tested on participants from the same cohort. In contrast, the same exPGS achieved slightly lower correlation coefficients when tested on a different cohort (0.04 ∼ 0.07 for four tissues individually and 0.08 for all tissues combined for UKB participants, P < 2.2×10^-16^). A reverse analysis using the genes detected in the UKB genes on GERA BP traits achieved a similar, but consistently better, correlation probably due to the larger gene sets in the exPGS models. We sought to further improve the exPGS models by introducing LASSO regression as implemented by GLMnet,^25^ which allows feature (gene) selection by cross-validation. This reduced the number of genes included in each model by removing correlated genes, but its optimal combination of genes resulted in an increased performance. We refer to these models as LASSO-exPGS. The combined all-tissue LASSO-exPGS models were created as before, with the same gene appearing up to four times if present in multiple tissues. A feature is defined as a gene in a specific tissue (gene: tissue), thus enabling a gene to be selected from more than one tissue in the all-tissue model, but, of course, from different CREs. This is because a gene may have different effects in different tissues. We found that the LASSO-exPGS achieved a consistently higher correlation with the phenotype. The correlation was higher by up to 10% in single tissue models, but 50%-60% higher in the combined tissue model, despite having a smaller number of genes in each model.

**Figure 5:**
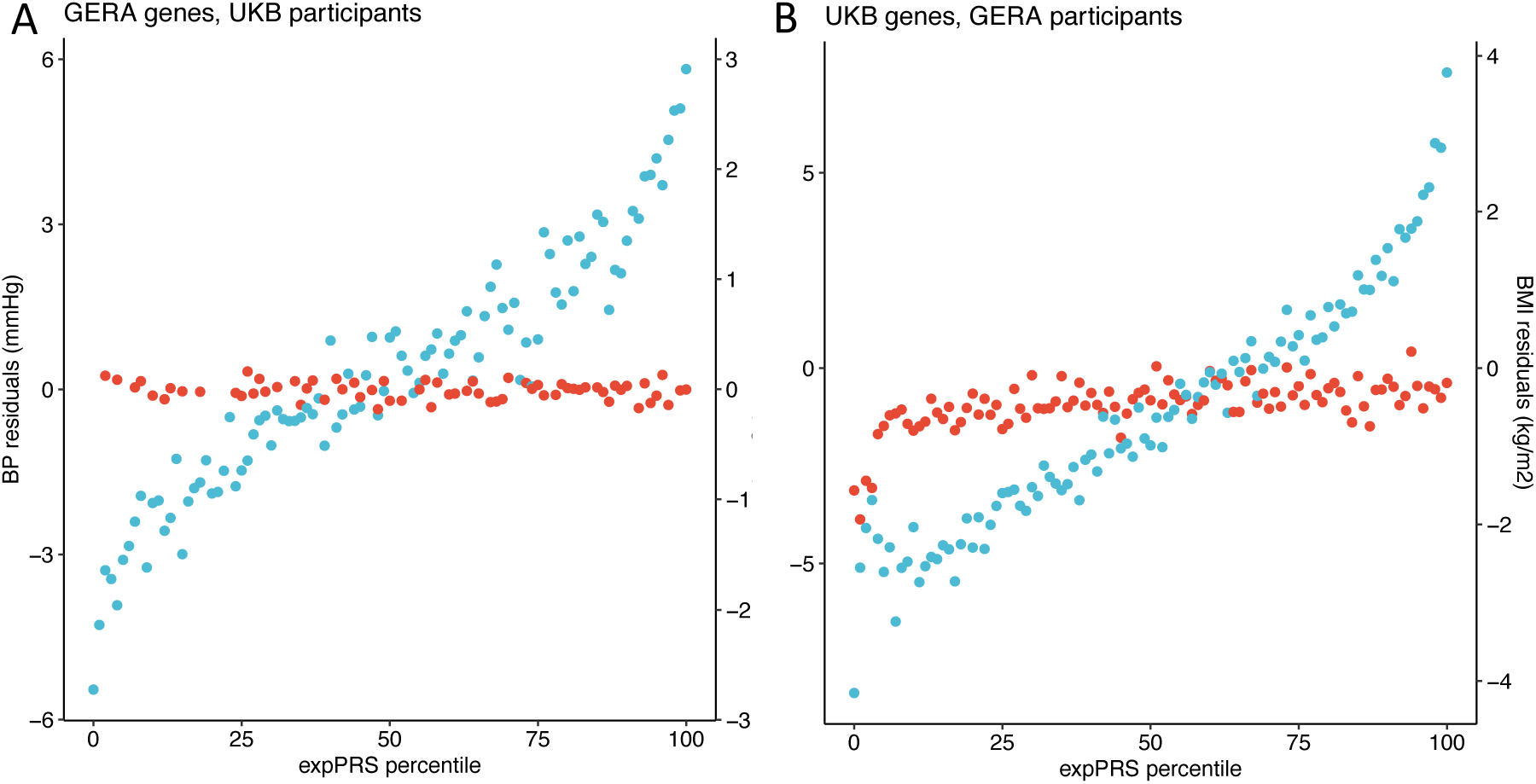
Performance of the expression polygenic score (exPGS) in the GERA and UKB cohorts for SBP (blue, test trait) and BMI (red, control trait). **A.** We show the combined all-tissue exPGS based on SBP genes detected in the GERA SKAT-CRE analysis but applied to UKB participants’ predicted expression for SBP and BMI, adjusted for age and sex. **B.** We show the complementary analysis of combined all-tissue exPGS based on SBP genes detected in the UKB SKAT-CRE analysis but applied to GERA participants’ predicted expression for SBP and BMI, adjusted for age and sex.

**Table 1:** Pearson correlation coefficients (R) between the expression polygenic score (exPGS) and LASSO-exPGS for SBP. For LASSO-exPGS models, the number of genes in the final model is in parenthesis. All results are statistically significant at P < 2.2×10^-16^.

|  | GERA participants |  |  |  | UKB participants |  |  |  |
| --- | --- | --- | --- | --- | --- | --- | --- | --- |
|  | GERA genes |  | UKB genes |  | GERA genes |  | UKB genes |  |
|  | exPGS | LASSO-exPGS | exPGS | LASSO-exPGS | exPGS | LASSO-exPGS | exPGS | LASSO-exPGS |
| Adrenal | 0.09 | 0.10<br>(n=127) | 0.14 | 0.16<br>(n=432) | 0.06 | 0.07<br>(n=142) | 0.13 | 0.15<br>(n=1,017) |
| Heart | 0.09 | 0.10<br>(n=130) | 0.15 | 0.17<br>(n=526) | 0.06 | 0.07<br>(n=109) | 0.13 | 0.16<br>(n=1,091) |
| Kidney | 0.06 | 0.07<br>(n=74) | 0.14 | 0.11<br>(n=208) | 0.04 | 0.04<br>(n=70) | 0.09 | 0.11<br>(n=623) |
| Artery | 0.09 | 0.11<br>(n=159) | 0.16 | 0.19<br>(n=613) | 0.07 | 0.08<br>(n=146) | 0.15 | 0.18<br>(n=1,272) |
| All tissues combined | 0.10 | 0.15<br>(n=335) | 0.19 | 0.24<br>(n=988) | 0.08 | 0.10<br>(n= 383) | 0.16 | 0.24<br>(n=2,875) |

Importantly, we achieved the best Pearson correlation coefficients for the LASSO-exPGS models using UKB genes for all tissues as 0.24 (P < 2.2×10^-16^) in both cohorts (**Table 1**). This result is significant as it explains a large proportion of the SNV-wide heritability (h^2^). Since the reported^4^ heritability for SBP is 0.213, our model explains ∼27% (0.24^2^/0.213) of the SNV-wide heritability.

### Gene Function and Validation

Genome-wide significant (P<5×10^-8^) associations from a prior GWAS (Evangelou^4^, GWAS catalog^23^) map to 3,283 SBP and 3,493 DBP genes. Of these, roughly half were also significant in our SKAT-CRE analysis in at least one of the four tissues tested, demonstrating both genetic variant-BP association as well as evidence that variants only within CREs of the putative gene showed BP association in target tissues. We further narrowed the list of putative genes by testing association between the predicted gene expression of putative genes with BP in the same tissue where the CRE associations were detected. This allowed us to explain genome-wide BP associations in terms of their CREs and downstream gene expression BP associations in the same tissue. A total of 598 SBP and 594 DBP genes passed these tests at a FDR<0.05. These genes have the greatest evidence of causality from three distinct statistical tests. As an example, consider the adjacent genes *MTHFR* and *NPPB*. **Figure 6A** shows the Locus Zoom plot of SBP GWAS data (Evangelou et. al^4^) for these two genes. As can be observed, the genome wide significant hits are close to both genes and it is impossible to determine which gene is causal, if any, and in which tissue. From predicted tissue gene expression, **Figure 6B** shows that *NPPB* has CRE supporting evidence in the heart, together with downstream predicted expression evidence in the heart, but not in any of the other three tissues. This is further supported by the known function of the gene, which encodes a protein involved in blood volume regulation, predominantly by the heart^26^. *MTHFR*, however, has supporting CRE evidence in multiple tissues, but its highest statistical significance is in the artery (**Figure 6C**). Clearly, our gene expression analysis is only significant in the artery and not in the three other tissues. This is consistent with previous work on the role of *MTHFR* in blood pressure regulation^27^ and with our previous work demonstrating significant CRE associations of *MTHFR* with BP in arteries^7^ and in arteries and heart^8^. These results provide the evidence to support causality of both *MTHFR* and *NPPB*, but with distinct biological functions in distinct tissues. Of the 598 SBP and 594 DBP genes that passed all steps of the validation analysis (**Figure 6D**), most of the genes could only be validated in one of the four tissues (**Supplementary Figure S9**). Only <25 genes were validated in all four tissues, for each of the traits (SBP and DBP). Due to the power differences between tissues in predicting gene expression (**Supplementary Figure S5**), it was expected that the overall validation analysis would detect a larger number of genes in tissues with high power (e.g., artery).

**Figure 6:**
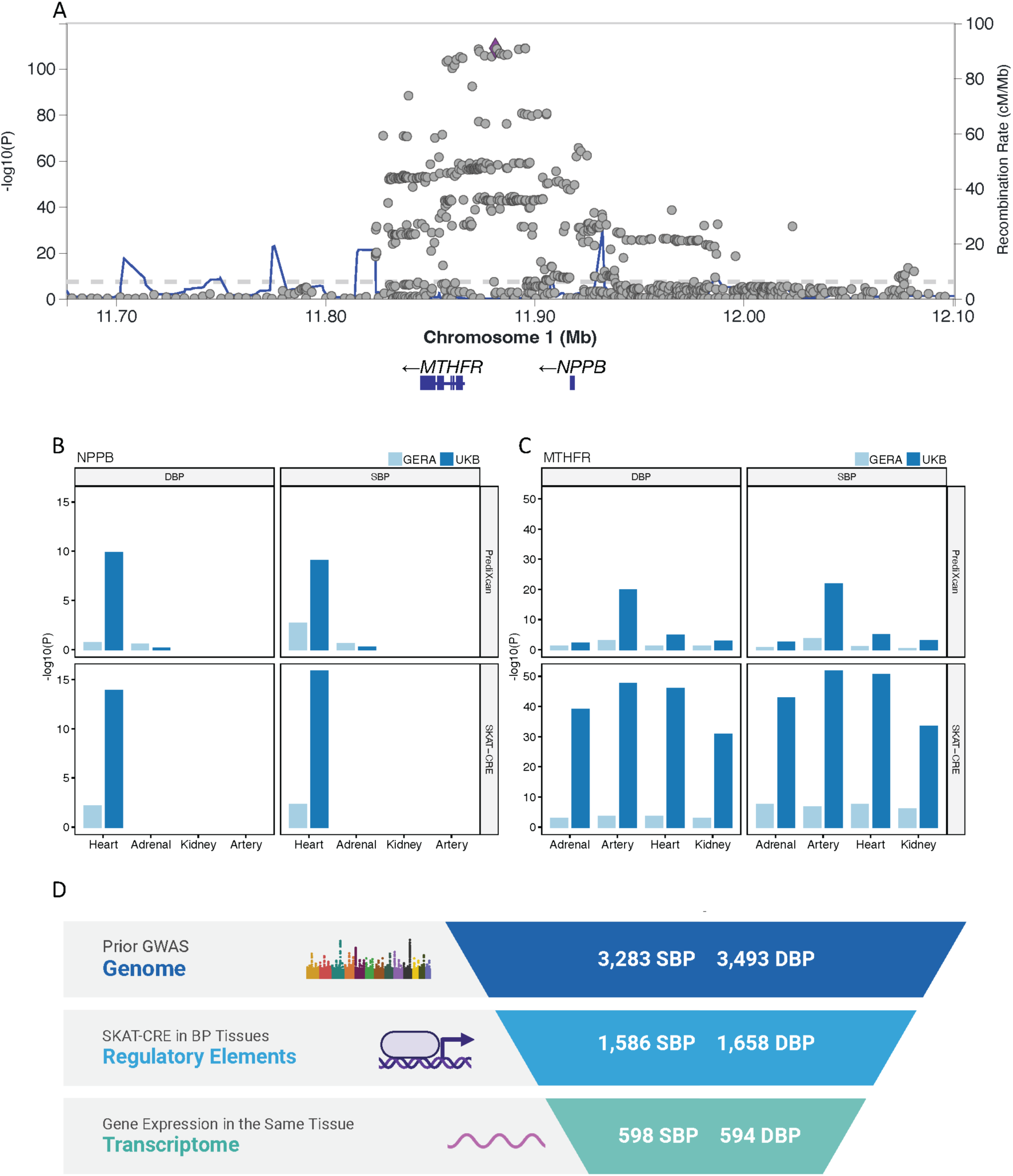
**A.** Locus Zoom plot of SBP GWAS summary statistics from the Evangelou et al. study^4^. **B.** and **C.** Associations of SBP and DBP with CREs (SKAT-CRE) and gene expression (PrediXcan) in specific tissues in both the UKB and GERA cohorts. Missing data in a tissue are from genes with TPM<1 for that tissue. **D.** Number of genes with statistically significant associations validated in each step of the analysis. Starting with genes associated with SBP and DBP in GWAS analysis, the subset of these that have significant SKAT-CRE associations with BP in specific tissues, and the subset of these genes with significant gene expression association with BP in the same tissue as the SKAT-CRE association.

## DISCUSSION

In this study, we discovered 3,220 SBP and 3,073 DBP-related genes in the adrenal gland, heart left-ventricle, kidney, and tibial artery, half of which were suggested from prior GWAS mapping and half of which, 1,634 SBP and 1,415 DBP genes, are novel candidates with respect to BP regulation. By constructing expression polygenic risk scores using only 988-2,875 of these genes, we explain ∼27% of the reported SNV-wide heritability (h^2^). This is substantially greater than that expected based on our previous work^8^, which estimated the partitioned heritability for ∼15,000 genes in these tissues to explain 40-50% of heritability. Here we explain more than half of our previously reported tissue partitioned heritability, by including only a fraction of the ∼15,000 genes, demonstrating that these genes are enriched for BP heritability. Just as the veracity of GWAS associations arises from statistical replication in an independent study, so do the genes identified here. These results arise from the novelty of our methods in which we restrict attention to the functionally relevant noncoding universe of CREs on a tissue-by-tissue basis, and demonstrate in individual subjects that variants within these CREs are associated with their measured BP, as well as associated with their tissue-predicted gene expression of that gene, and, that the cumulative effects of all their gene expression changes are correlated with their BP. Therefore, this is a first demonstration, for BP specifically but for any polygenic trait generally, of the veracity of the now-standard hypothesis that noncoding variants associated with a complex trait are regulatory and affect the trait through gene expression changes in many genes across the genome in different tissues. Our results are specific and, therefore, subject to experimental tests.

The magnitude of the universe of functional elements, variants affecting CRE function, numbers of CREs per tissue and the target genes, is still not fully known. So far, in this study, each protein-coding gene has a median of 10-20 CREs in each tissue, and each gene included a median of 70-100 CRE variants for ∼12,000 genes that are expressed in each tissue. In other words, ∼3-7 regulatory variants per gene per tissue affect BP. The inter-individual BP variation explained by these combined observations is highly significant (r= 0.24, P < 2.2×10^-16^) but there are additional molecular details to be identified. There is considerable evidence that many of these BP genes are pleiotropic, across the four tissues studied (**Figure 2**). However, it is unknown whether this expression pleiotropy arises from shared or tissue-restricted CREs, as our recent research shows.^8^ Thus, even for the same gene, its genetic effect on BP may arise from different variants and CREs in distinct tissues, adding a level of complexity not envisioned before, but testable. Additionally, where is the remaining unexplained SNV heritability? These will likely come from SNVs of lower frequency which recently have been demonstrated to make substantial contributions to complex trait heritability.^28^ Importantly, there are other tissues that are also likely involved in BP regulation, such as the brain.^29^

The tissue contributions to a polygenic trait, particularly a physiologically significant trait like BP, is an important direction for genome analysis. As epigenomic maps of multiple tissues emerge,^30^ the methods we have developed, previously published and here, will allow us to scan each tissue for their contribution to BP variation (**Table 1**).^8^ Nevertheless, so far, in both cohorts, we found that for BP the arterial effect is greater than that of other tissues and the renal contribution the least. This finding is consistent with our recent^8^ and previous work.^9,10^ However, it is important to note that paucity of high-quality chromatin assay and gene expression data on a specific tissue may underestimate its contribution. Thus, yet better and comprehensive epigenomic maps are necessary.

Finally, this work provides information on specific tissues based on bulk epigenomic assays and does not discriminate between cell types and tissue structures. The tissues we examined comprise functionally diverse cell types; assessing their contribution is crucial to understanding the genetic component to BP physiology. Our next challenge is to use single cell data to provide this greater resolution of BP genomic biology by identifying cell type-specific CREs, their target genes’ expression and their quantitative contributions to BP variation.^30^

Our underlying working model is that for each tissue, only some variants in CREs active in that tissue alter target gene expression in conjunction with other CREs of that gene, and, that an ensemble of such genes in that tissue affect BP by altering some yet unknown tissue phenotype. Therefore, despite the above advances, some major questions remain unanswered. First, although we can identify CREs active within tissues, including those with high-impact variants, how do we assign each such CRE to its target gene? Even with that answered for specific CRE variants, such as with Hi-C or other genomic proximity ligation data,^20,31^ how can we demonstrate that the *ensemble* of identified CREs for specific genes explain its gene expression variation in that tissue? Finally, is the expression variation of such tissue-wise genes a significant explanation of BP phenotypic variation? These studies can be further improved because a statistical model of the epigenome of a tissue can be used to identify additional CREs missed in an experimental assay, predict the effect of all sequence changes within CREs as well as estimate the contribution of specific common or rare CRE variants to BP regulation. Because these genotype-phenotype data are readily available for biobank participants, tissue-based CRE prediction can augment and add an additional dimension to living-subject biobanks to provide hypotheses that can be tested in experimental paradigms.

A potential limitation of our work is that although these findings were significant in our statistical analysis, we noticed that the SKAT-CRE test statistics were inflated, as seen in the quantile-quantile plots (**Supplementary Figure S2**). We believe that the null distribution used by the SKAT test is overly naïve. Nonetheless, we were able to replicate 50%-70% of the findings in an independent dataset (**Figure 3**), which far exceeded the chance replication rate of 12%-15%, as demonstrated by permutation analysis. In addition, we provided further supporting evidence through expression-based analysis.

## METHODS

### Study cohorts

We used two population cohorts in this study. The first was a ‘discovery’ cohort, the Genetic Epidemiology Research on Adult Health and Aging (GERA), which our previous work^9^ demonstrated had significant statistical power from its longitudinal BP records for identifying genomic variation underlying BP traits. This was owing to long-term averaging of noisy BP traits varying by extrinsic factors, such as stress, time of day, measurement instruments (manual vs. machine) and location of measurement. We studied 77,822 unrelated participants of self-described European ancestry, with directly measured or imputed genotypes, as previously described.^9^ Systolic (SBP) and diastolic (DBP) BP were studied. Second, we used the UK Biobank (UKB),^14^ a large population-based prospective study, as the ‘replication’ cohort; although it has few (∼2) BP measurements per individual, a much larger number of 315,270 unrelated participants of self-described British ancestry were examined. UKB genotype data were preprocessed and imputed by the UKB consortium as described in their online genome-wide SNP array-based genotyping documentation.^32^ Genotype imputation was performed using haplotypes from 2 different reference panels. The Haplotype Reference Consortium (HRC) was used first, since the HRC misses many sites observed in the 1000 Genomes Project;^17^ a further round of imputation used the UK10K^33^ and 1000 Genomes^17^ combined reference panels.

### BP phenotypes and covariate adjustments

A detailed description of the GERA phenotype processing is available in our previous publication.^9^ BP measurements in both cohorts were adjusted for age, age^2^, BMI and sex, with regression residuals used as the phenotype. Control phenotypes of height and BMI were only adjusted for sex and age. We added 15 mmHg and 10 mmHg to SBP and DBP values, respectively, to individuals treated with antihypertensives, as is standard in BP epidemiology and as used in our previous study.^9^

### Constructing comprehensive epigenomic maps from genome-wide chromatin accessibility data

We have previously published machine-learning methods for constructing comprehensive enhancer (CRE) maps, which we used with minor modifications.^11–13^ Briefly, MACS2^34^ was used for peak calling while ATAC-seq samples were adjusted at the cut-sites by +4bp for the forward and -5bp for the reverse strand to account for the 9bp insertion by the Tn5 enzyme.^35^ We used chromatin accessibility data from publicly available ENCODE^20^ DNase-seq as well as in-house generated kidney ATAC-seq data, the latter occasioned by the limited availability of high-quality human kidney open chromatin data in public databases. In-house snap-frozen kidney tissue samples were procured from the Gift of Life Michigan and The National Disease Research Interchange (NDRI). Detailed information on the ATAC-seq protocol used has been previously published.^8^ These kidney ATAC-seq data are available in the NCBI Gene Expression Omnibus (GEO)^36^ under accession number GSE200047.

### Sequence-based models for regulatory variant prediction

For identifying sequence variants with regulatory effects, we built gkm-SVM (machine learning) models as described in our previous work.^12,13^ Briefly, after quality control analyses, for each high-quality sample, we defined tissue-specific open chromatin regions as a positive training set and then removed elements with >1% of N-bases, >70% of repeats, and common (active in at least 30% of samples across all ENCODE data sets) open regions, as previously described.^11^ We only used regions that also overlapped H3K27ac peaks from the same tissue and used 600bp fixed-length regions as a training set by extending ±300bp from each peak summit. As a negative training set, we randomly selected an equal number of genomic regions matching length, GC content, and sequence repeat fraction as in the positive set. We used LS-GKM^13^ for training with l=11, k=7, d=3 and t=4 (weighted-gkm kernels). For each sample, we generated ten different models with different random samplings of negative training sets, and further combined these models to generate one model per tissue. These models were used to estimate the impact of each SNV within each CRE by calculating deltaSVM^18^ scores (see Methods) for all common (minor allele frequency (MAF) > 1% in the 1000 Genomes^17^ v3 data) SNVs in the CRE map, for each tissue. We used ±10 bp regions centered on these variants for scoring. The training sets used and the final models generated are available in our previous work.^11^

### SKAT-CRE test

SKAT association tests were performed using the SKAT package for R v1.3.2.1 and genotypes were loaded using Seqminar v8.0. The phenotypes used were adjusted SBP and DBP. Only individuals with self-reported European ancestry were included in both GERA and the UKB cohorts. Related individuals, with kinship coefficients greater than 0.088 signifying third degree or closer relatives, as estimated using the KING software as implemented in PLINK 2.o,^37^ were excluded using a greedy algorithm in PLINK to minimize the number of excluded samples. Genotype imputation and processing was as previously described.^9^

For analysis, genes were defined as the gene body as in GENCODE V19^20^ with an additional 50kb on either side of the gene start and end, including all introns. We assigned the absolute value of the deltaSVM value for each variant as weights in SKAT analysis. Adjustments for multiple comparisons used the Benjamini-Hochberg method in R with a false discovery rate (FDR) of 5%.

### PrediXcan analysis

We constructed our own PrediXcan models for CRE-restricted and all-variant models, following the recommended PredictDB pipeline.^38^ We used GTEx v8,^21^ and lifted over the genotype data to GRCh37 using Picard v2.18.11. PrediXcan models are considered statistically significant if their mean Pearson correlation coefficient exceeds 0.1 across cross-validation, with estimated p≤0.05, as determined by the PredictDB pipeline.^38^ The PredictDB pipeline repository was downloaded from GitHub on August 30, 2021. We used PrediXcan,^24^ as included in the MetaXcan v0.7.4, to predict target gene expression using statistically significant models. We then used PrediXcan to estimate the association of its predicted expression with SBP and DBP. Multiple comparisons adjustment was performed using the Benjamini-Hochberg method in R, FDR < 5%.

### Statistical and computational analysis

All statistical and computational analyses were performed in R v3.5.1. Additional R packages used were SKAT v1.3.2.1, Seqminer v8.0, GLMnet v2.0 and data-table v1.12.8. The graphical plots were generated using ggplot2 v3.3.2 and ggpubr v0.2.4. LocusZoom plot was created using LocusZoom^39^. Upset plots were created in python (3.7.1) using the upsetplot library v0.8.0. Illustrations were created on BioRender^40^ under academic license. Python 3.7.1 was used for PredictDB and MetaXcan (PrediXcan). General operations on genotype files were performed using bcftools v1.11, qctool v2.0.5, PLINK v1.9 and v2.0, htslib 1.9, and Picard v2.18.11. Permutation analysis was performed by random sampling without replacement, repeated 100,00 times.

## Data Availability

1. Raw sequencing data is publicly available from our recent publication (PMID: 37910504, DOI: 10.1016/j.celrep.2023.113351) on NCBI Gene Expression Omnibus (GEO: GSE200047, see link)
2. All data produced in the study are available on OSA (See link).

https://osf.io/54tsf/?view_only=e499ebb845054b999b432216050c3369)

https://www.ncbi.nlm.nih.gov/geo/query/acc.cgi?acc=GSE200047

## SUPPLEMENTARY MATERIAL

Supplementary Figures (.docx file).

SKAT-CRE results Table, UKB and GERA (.csv file).

PrediXcan results Table, UKB and GERA (.csv file).

Regulatory maps per tissue, hg19 (4 .bed files).

Files are available on OSF https://osf.io/54tsf/?view_only=e499ebb845054b999b432216050c3369

