## Supplemental Figures for "Identification of the molecular components of enhancer-mediated gene expression variation in multiple tissues regulating blood pressure"

Supplementary Figure S1:

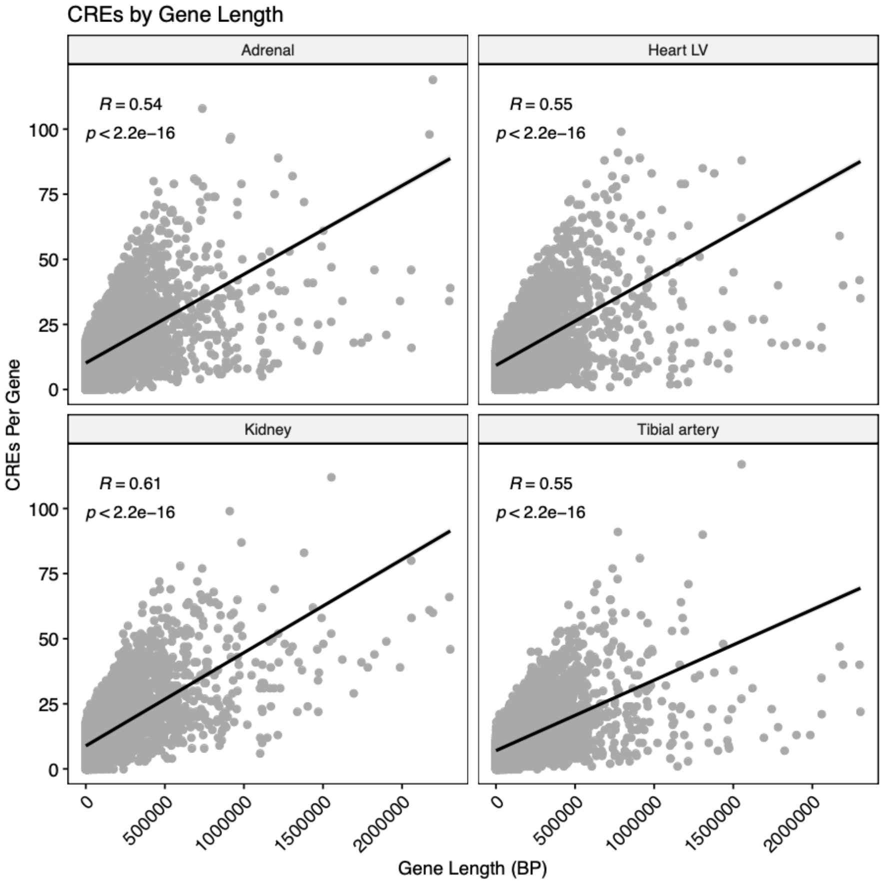


**Figure S1:** Numbers of CREs per gene is proportional to gene length. The Y-axis shows the number of CREs for each gene and the X-axis is the gene length in base pairs (bp). The best fitting regression line, the Pearson correlation coefficient and significance value are provided for each tissue.

Supplementary Figure S2:

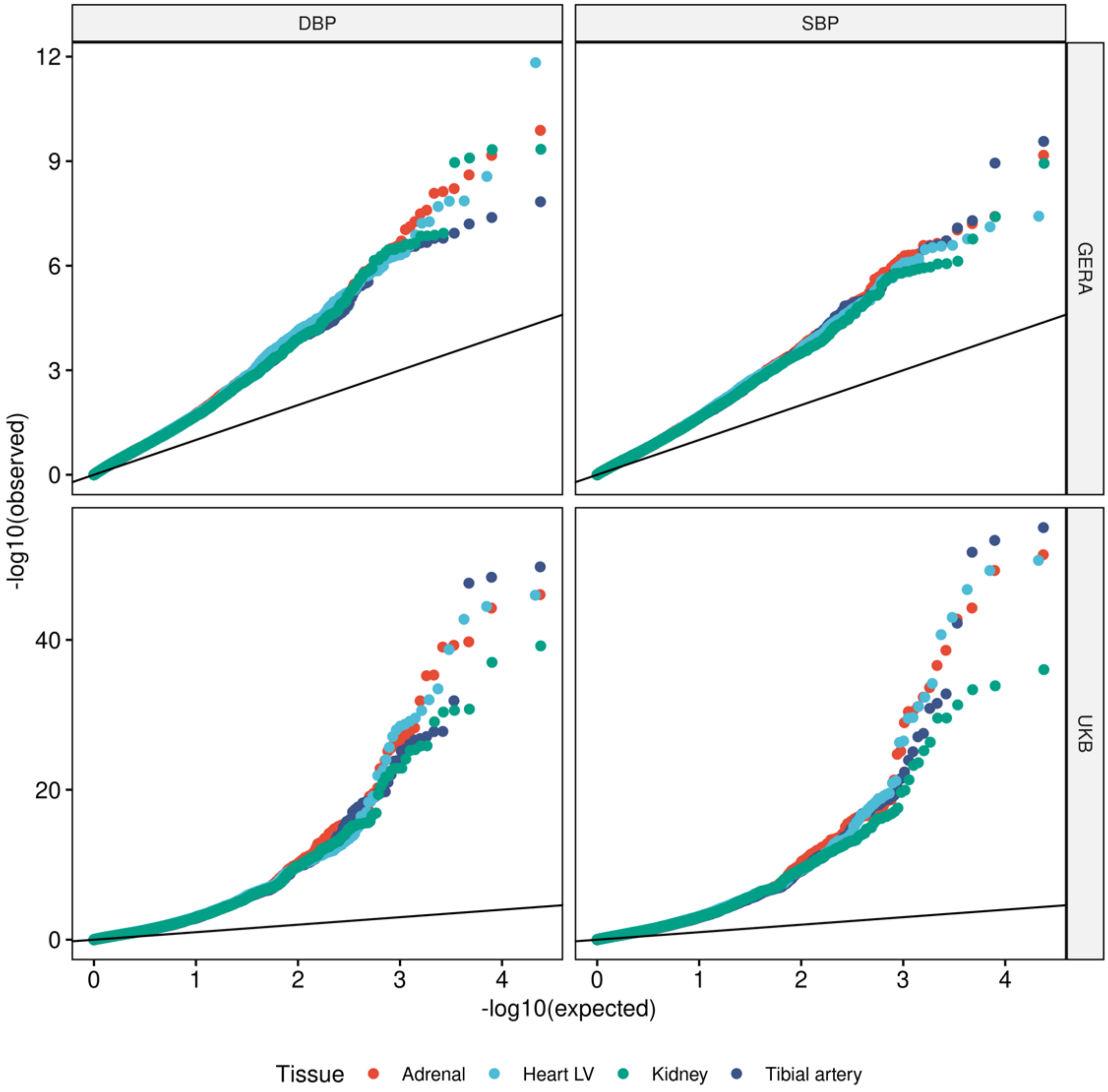


**Figure S2:** Quantile-quantile plots of expected and observed -$\log_{10}$p values of the SKAT-CRE test statistic for SBP and DBP. The top and bottom panels represent the GERA and the UKB cohorts, respectively; the right and left panels represent SBP and DBP associations, respectively. Each tissue is represented by a different color.

Supplementary Figure S3:

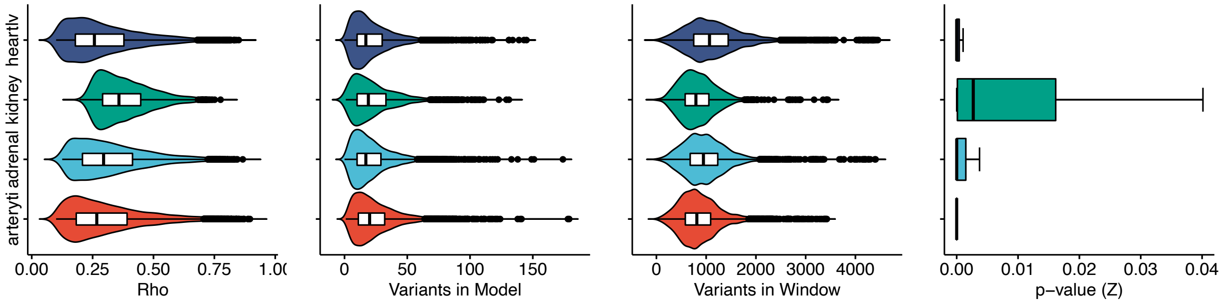


**Figure S3:** Statistical properties of CRE-restricted LASSO regression models. The different tissues are represented by different colors and labeled so on the Y-axis. The panels represent (from left to right) the Pearson correlation of predicted expression in cross validation analyses, the number of variants selected in each model, the number of variants available to the algorithm for creating each model, and the significance levels from cross-validation.

Supplementary Figure S4:

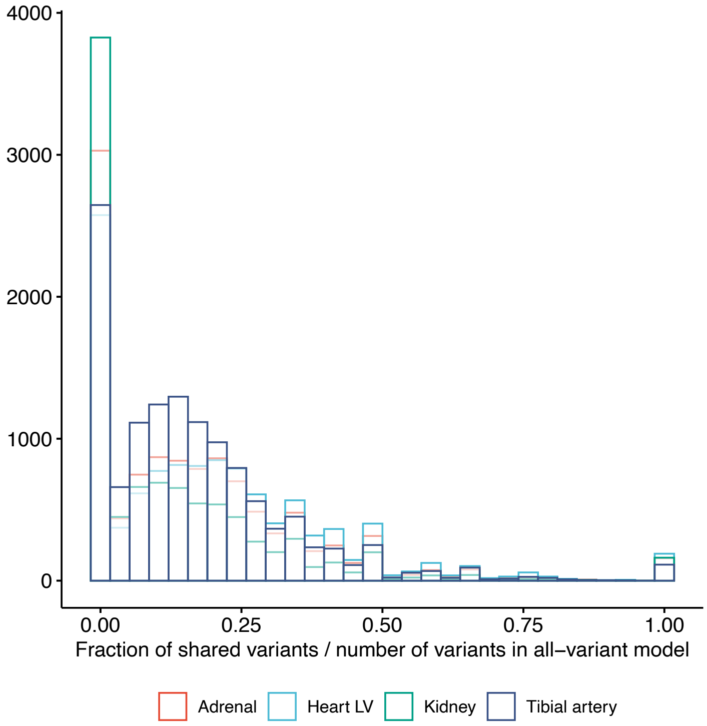


**Figure S4:** Overlap between variants in the CRE-restricted and all-variant models. The overlap is represented as the fraction of shared variants out of the number of variants in the all-variant model, which is always higher.

Supplementary Figure S5:

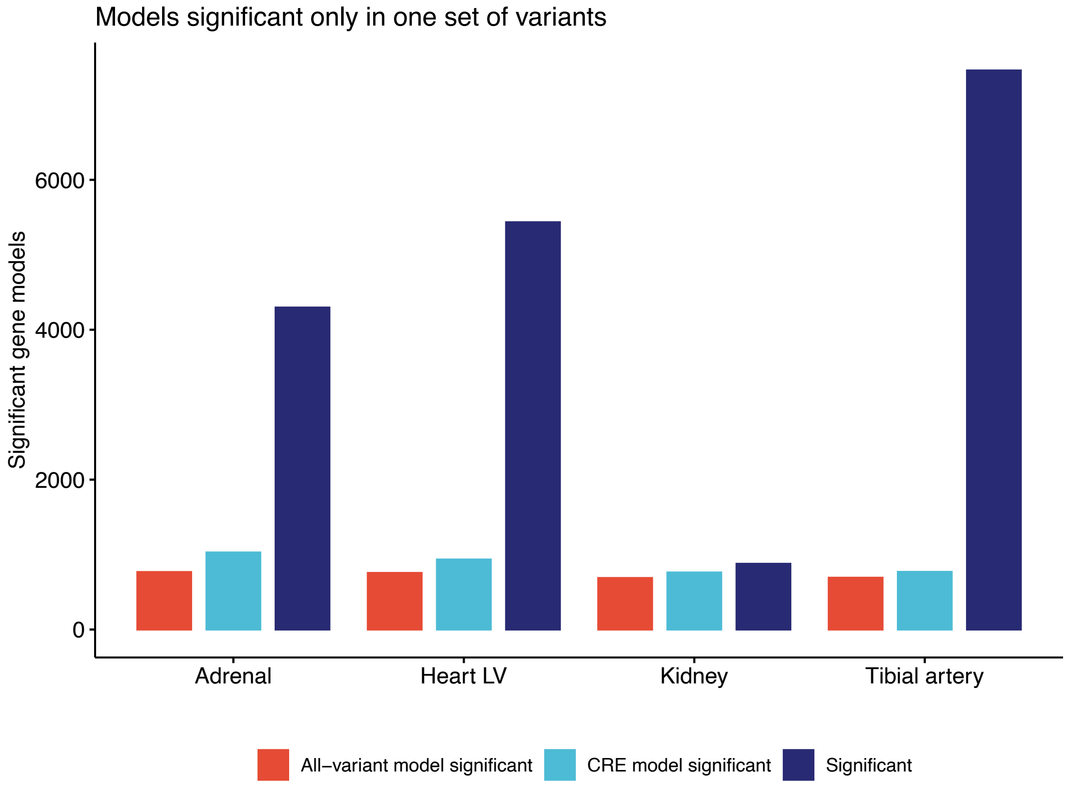


**Figure S5:** Statistical properties of gene models significant for gene expression prediction. A comparison between CRE-restricted and all-variant models with the bars representing the number of gene models that are significant only in the all-variant models (red), only in the CRE-restricted models (light-blue) or in both (dark-blue).

Supplementary Figure S6:

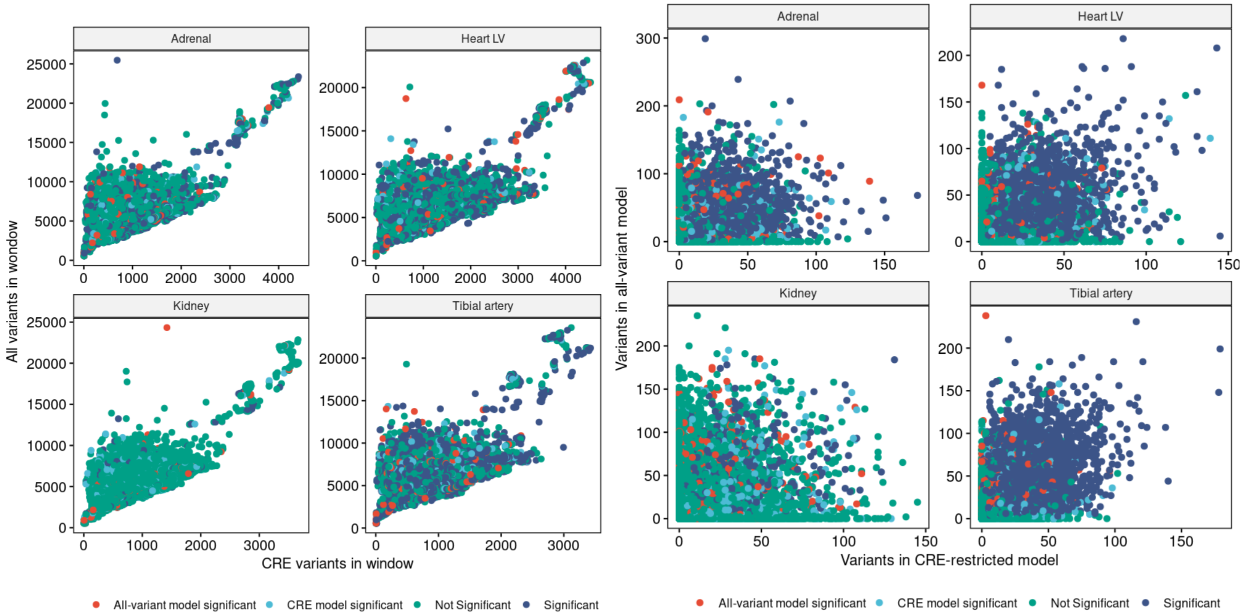


**Figure S6:** Numbers of variants available versus those selected in the final model. The plots compare all-variant (Y-axis) and CRE-restricted (X-axis) models with each panel representing a different tissue. Models were labelled as those that were significant in both (dark-blue), only in the CRE-restricted (light-blue), only in the all-variant (red), and significant in neither (green) models.

Supplementary Figure S7:

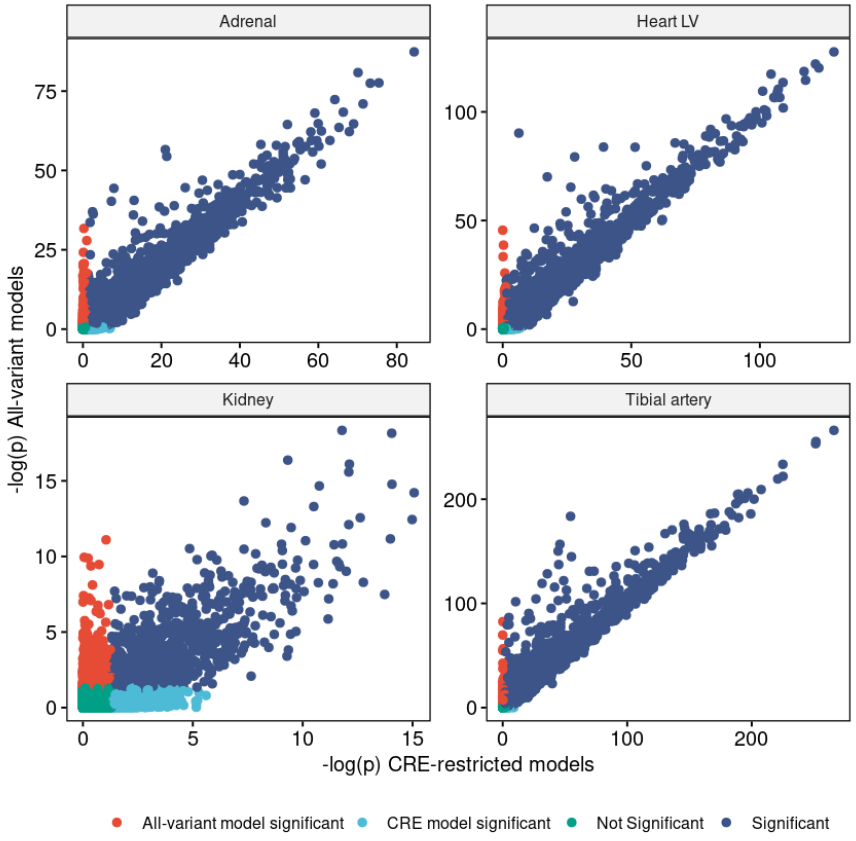


**Figure S7:** Comparison of significance values (-log$_{10}$p) between CRE-restricted and all-variant models. PredictDB model significance is dependent on Pearson correlation during cross validation in addition to significance levels. Models are labelled as those that were significant in both (dark-blue), only in the CRE-restricted (light-blue), only in the all-variant (red), and significant in neither (green) models.

Supplementary Figure S8:

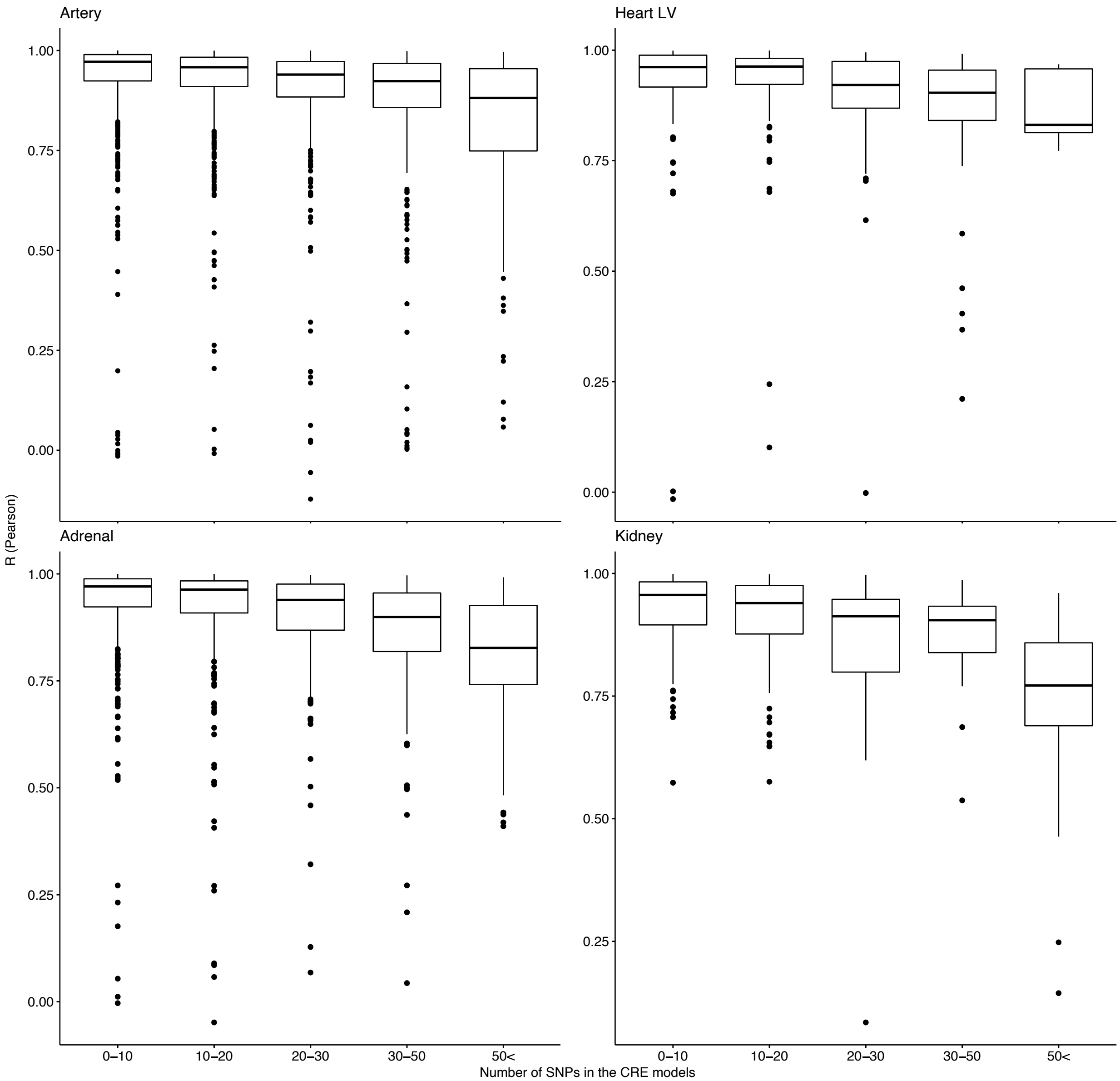


**Figure S8:** Gene expression prediction comparisons between all-variant and the CRE-restricted models, using the GERA cohort genotypes. Pearson correlations between the two predictions (Y-axis) are shown stratified by the numbers of variants in each CRE-restricted gene-model (X-axis).

Supplementary Figure S9:

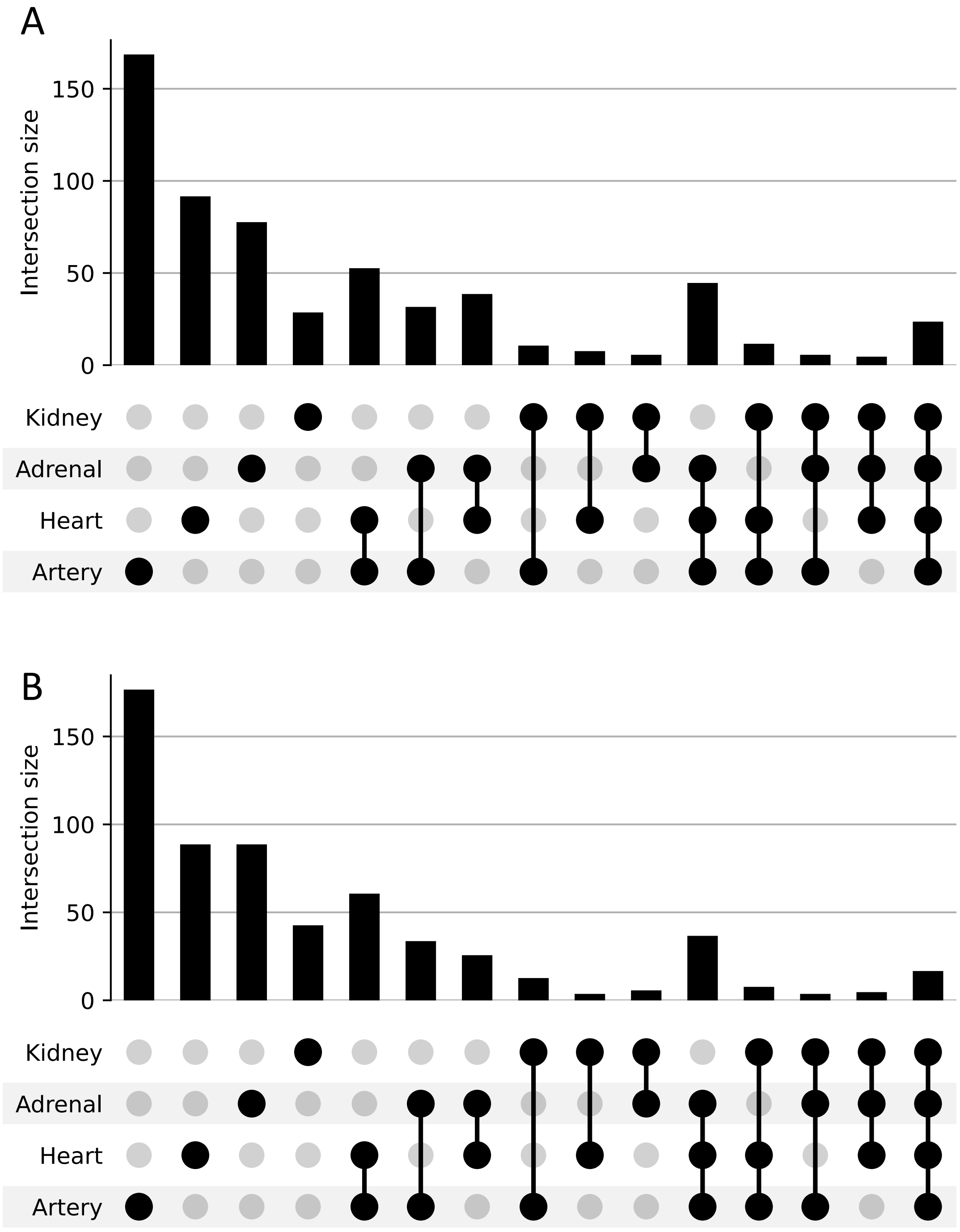


**Figure S9:** Numbers of **A.** DBP and **B.** SBP genes that passed all steps of the validation analysis, showing evidence of GWAS, SKAT-CRE and predicted expression significant associations with BP in each tissue, and the overlap between the different tissues.
